# Revisiting *Plasmodium vivax* molecular correction

**DOI:** 10.64898/2026.06.02.26354709

**Authors:** Aimee R Taylor, Yong See Foo, Michael T White

**Affiliations:** Institut Pasteur, Université Paris Cité; University of Melbourne

## Abstract

**Background:** Reliable inference of *Plasmodium vivax* recurrence states — relapse, recrudescence and reinfection (the “3Rs”) — improves estimates of antimalarial efficacy. The R package Pv3Rs features a Bayesian model designed for *P. vivax* molecular correction, i.e., using parasite genetic data to infer recurrence states. The model is an extension of a prototype built to analyse microsatellite data from the Vivax History (VHX) and Best Primaquine Dose (BPD) trials.

**Methods:** We re-analysed data from 212 VHX and BPD trial participants (493 recurrences) using Pv3Rs, comparing results with those from the prototype and with genetic relatedness estimated using Dcifer, a tool for estimating relatedness based on identity-by-descent. Posterior recurrence state probabilities were computed using both uniform and time-to-event priors: artificial but equal prior probabilities facilitate posterior interpretation, while time-to-event priors leverage all available information and enable re-computation of failure rates. Relatedness estimates were used to identify and correct instances of model misspecification.

**Results:** The Pv3Rs model generated posterior probabilities for all recurrences and was able to jointly model data on all episodes per participant for 89% of participants, compared with 73% using the prototype. Recurrence state probabilities were broadly consistent across methods, though the Pv3Rs model elevated reinfection probabilities slightly. Relatedness estimates exposed various outliers consistent with half-sibling parasites and/or genotyping errors. Outlier correction impacted some per-participant failure probabilities, but reinfection-adjusted radical-cure failure rates of high-dose primaquine remained near 3%, in line with previous findings.

**Conclusion:** Re-analysis of VHX and BPD *P. vivax* genetic data restates earlier reinfection-adjusted efficacy estimates. It demonstrates the increased computational capability and misspecification sensitivity of Pv3Rs, highlighting a need for careful analyses. Using relatedness-based diagnostics alongside model-based inference, we were able to harness the advantages of model-based inference and provide a framework for future *P. vivax* molecular correction.

## 1 Introduction

Of the two major causes of human malaria, *Plasmodium vivax* is the hardest to eliminate, owing largely to its ability to relapse via activation of dormant liver-stage parasites called hypnozoites (Markus, 2011). Failure to fully treat a blood-stage infection (recrudescence) and/or a new mosquito inoculation (reinfection) can also cause recurrent blood-stage infection. Estimates of the source of recurrence can be used to improve estimates of antimalarial efficacy. For example, reinfection should be separated from relapse and recrudescence when estimating the efficacy of radical cure, i.e., treatment of both liver and blood-stage parasites.

Molecular correction uses parasite genetic data to estimate the source of recurrence and thus correct efficacy estimates. It is most developed for *P. falciparum*, where recrudescence and reinfection cause recurrence. Recrudescent parasites are asexual clones of parasites within infections that temporarily drop below the limit of detection following treatment. They are necessarily identical to pre-recurrence blood-stage parasites, aside from de novo mutations. Parasites derived from a new mosquito inoculation (reinfection) are related to earlier parasites only insofar as parasites among different mosquitoes within the surrounding population are related. The World Health Organisation (WHO) provides guidelines for *P. falciparum* molecular correction (World Health Organization, 2008, 2021). They are based on counting alleles that match across episodes. Alternatively, recurrence states can be inferred statistically, using either a probabilistic model (Plucinski et al., 2015; Mehra et al., 2025; Gerlovina et al., 2025), or a hypothesis test, where the null distribution is approximated by an empirical distribution of relatedness between parasites sampled from different people (Plucinski and Barratt, 2021).

In addition to recrudescence and reinfection, *P. vivax* molecular correction contends with relapse. Relapsing parasites derive from hypnozoites, which are draws from one or more past inoculations. They are related to earlier parasites insofar as parasites are related between and within mosquitoes. An obligate stage of sexual recombination occurs within mosquitoes, with self- and/or cross-fertilisation between one or more parasite pairs, generating offspring that are perfectly related clones, relatively unrelated strangers and partially related siblings, including meiotic siblings (reciprocal siblings derived from a single cross-fertilised gamete pair), parent-child-like siblings (derived from two gamete pairs: one self-fertilised, the other cross-fertilised), regular full siblings (derived from two gamete pairs: both cross-fertilised with the same two genotypes) and half siblings (derived from two gamete pairs: both cross-fertilised with only one common genotype); see Figure S1 of Foo et al. (2026). Accordingly, relapsing parasites include clones, strangers and siblings (Popovici et al., 2018, 2026), with evidence of meiotic siblings (Bright et al., 2014). Unsurprisingly, given this complexity, there are no WHO guidelines for *P. vivax* molecular correction. Conventionally, *P. vivax* recurrences are classified using descriptive summaries of genetic similarity between episodes, e.g., as genetically heterologous or homologous (Imwong et al., 2007, 2012). Increasingly, newer studies feature model-based estimates of genetic relatedness (Kleinecke et al., 2025; Kambuaya et al., 2025; Rosado et al., 2026; Tadesse et al., 2026; Popovici et al., 2026).

White et al. (2018) first attempted to model per-recurrence probabilities of relapse, uncovering a need for multi-locus genetic data. Not long after, Taylor et al. (2019) demonstrated the feasibility of statistical genetic inference of *P. vivax* recurrence states using data on ≤ 9 microsatellites. Posterior probabilities of relapse, reinfection and recrudescence (3Rs) were computed under a Bayesian model, summing over graphs of clonal, sibling, and stranger relationships between inter- and intra-episode parasites. All graphs are compatible with relapse, while only a subset with inter-episode clonal/stranger edges are compatible with recrudescence/reinfection. The model was fit to data from two clinical trials, the Vivax History (VHX) and Best Primaquine Dose (BPD) trials (Chu et al., 2018, 2019). Using posterior probabilities, a reinfection-corrected failure rate of radical cure with high-dose primaquine (7 mg/kg total dose) was calculated. At ~ 3%, it was significantly lower than reinfection-uncorrected failure rate (12%, recurrence in 80/654 patients) reported previously (Chu et al., 2019), and is broadly consistent with the recurrence rate of 5% (2 of 43 participants) reported among participants receiving high-dose primaquine in a Cambodian clinical trial where relocation of participants prevented reinfection (Eng et al., 2025).

In this study, we revisit *P. vivax* molecular correction for the VHX and BPD trials using a new R package Pv3Rs (Foo et al., 2026). Pv3Rs features an extension of the prototypic model of Taylor et al. (2019). Pv3Rs also supports visual inference via a data-plotting function. We compare results with those from the prototype and with analyses of relatedness estimated using Dcifer (Gerlovina et al., 2022). To assess how model modifications affect previously reported failure rates, we recompute posterior probabilities using the time-to-event prior probabilities reported in Taylor et al. (2019). To highlight differences between the prototype, Pv3Rs, and relatedness-based analyses, posteriors are also computed using genetic information only, assuming recurrence states are equally likely *a priori*.

## 2 Methods

### 2.1 Clinical trials

The VHX (Chu et al., 2018) and BPD (Chu et al., 2019) trials were both conducted by the Shoklo Malaria Research Unit which operates clinics on the Thailand-Myanmar border, a region of low seasonal *P. vivax* transmission, with a population of migrant workers and displaced persons (Carrara et al., 2013). Microscopy was used to detect parasites (lower limit of detection approximately 50 parasites per *µ*L). Participants with uncomplicated *P. vivax* mono-specifies infection were randomised to different treatments including artesunate (AS), choloroquine (CQ), and dihydroartemisinin-piperaquine (DP), which kill blood-stage parasites only, and primaquine (PQ), which also kills hypnozoites. More specifically, 644 participants were randomised to receive AS (2 mg/kg/day for 5 days), CQ (25 mg base/kg over 3 days) or CQ plus PQ (7 mg base/kg over 14 days) in the VHX trial (May 2010 to October 2012). In the BPD trial (February 2012 to July 2014), 680 participants were randomised to 7 mg base/kg PQ over either 14 or 7 days and combined with either CQ or DP (7 mg/kg dihydroartemisinin and 55 mg/kg piperaquine over three days). Participants were followed up actively: daily until parasite negative, weekly for eight weeks in VHX and on weeks 1, 2, and 4 in BPD, and then monthly for a total of one year. Participants were encouraged to seek care between scheduled visits. However, fewer than 5% of recurrences were detected passively. All malaria-positive recurrences were treated under supervision, irrespective of symptoms. In VHX, CQ plus PQ over 14 days was given after 9 recurrences or earlier if clinically indicated; otherwise, recurrences were retreated as per randomisation. In BPD, recurrences were treated with CQ and PQ over 14 days. Both trials were approved by the Mahidol University Faculty of Tropical Medicine Ethics Committee (VHX: MUTM 2010-006; BPD: MUTM 2011-043, TMEC 11-008) and the Oxford Tropical Research Ethics Committee (VHX: OXTREC 04-10; BPD: OXTREC 17-11), and registered at ClinicalTrials.gov (VHX: NCT01074905; BPD: NCT01640574).

### 2.2 Microsatellite data

The previously published microsatellite data are described in detail in Taylor et al. (2019). In brief, microsatellite data on *P. vivax* parasites sampled from two or more episodes in the same study participant were generated successfully for 493 recurrences in 212 study participants (Figure S1). Among the 493 recurrences, 122 were in 109 participants treated with PQ (7 mg/kg total dose); 371 were in 103 study participants untreated with PQ (Taylor et al., 2019). Among the 212 participants, not all episodes detected microscopically were genotyped. Table 1 shows the distribution over the number of episodes genotyped per participant. Hereafter, participants are referred to by the trial in which they were enrolled and their per-trial unique identifier; an additional number preceded by an underscore marks each episode; e.g., VHX_419_6 corresponds to the sixth episode among all episodes (successfully genotyped or not) in VHX participant 419. Across all genotyped episodes, genotyping was attempted at markers `PV.3.27`, `PV.3.502` and `PV.ms8`. With some exceptions (e.g., VHX_457, Figure 1), if alleles match among a minority of successful attempts, no additional markers were typed; otherwise, six additional markers were typed. Not all markers typed were typed successfully.

**Table 1:** Numbers of episodes genotyped per participant.

|  |  |  |  |  |  |  |  |  |  |  |  |
| --- | --- | --- | --- | --- | --- | --- | --- | --- | --- | --- | --- |
| Number of episodes genotyped per participant | 2 | 3 | 4 | 5 | 6 | 7 | 8 | 9 | 10 | 11 | 14 |
| Number of participants | 131 | 29 | 8 | 12 | 6 | 5 | 8 | 6 | 5 | 1 | 1 |

**Figure 1.**
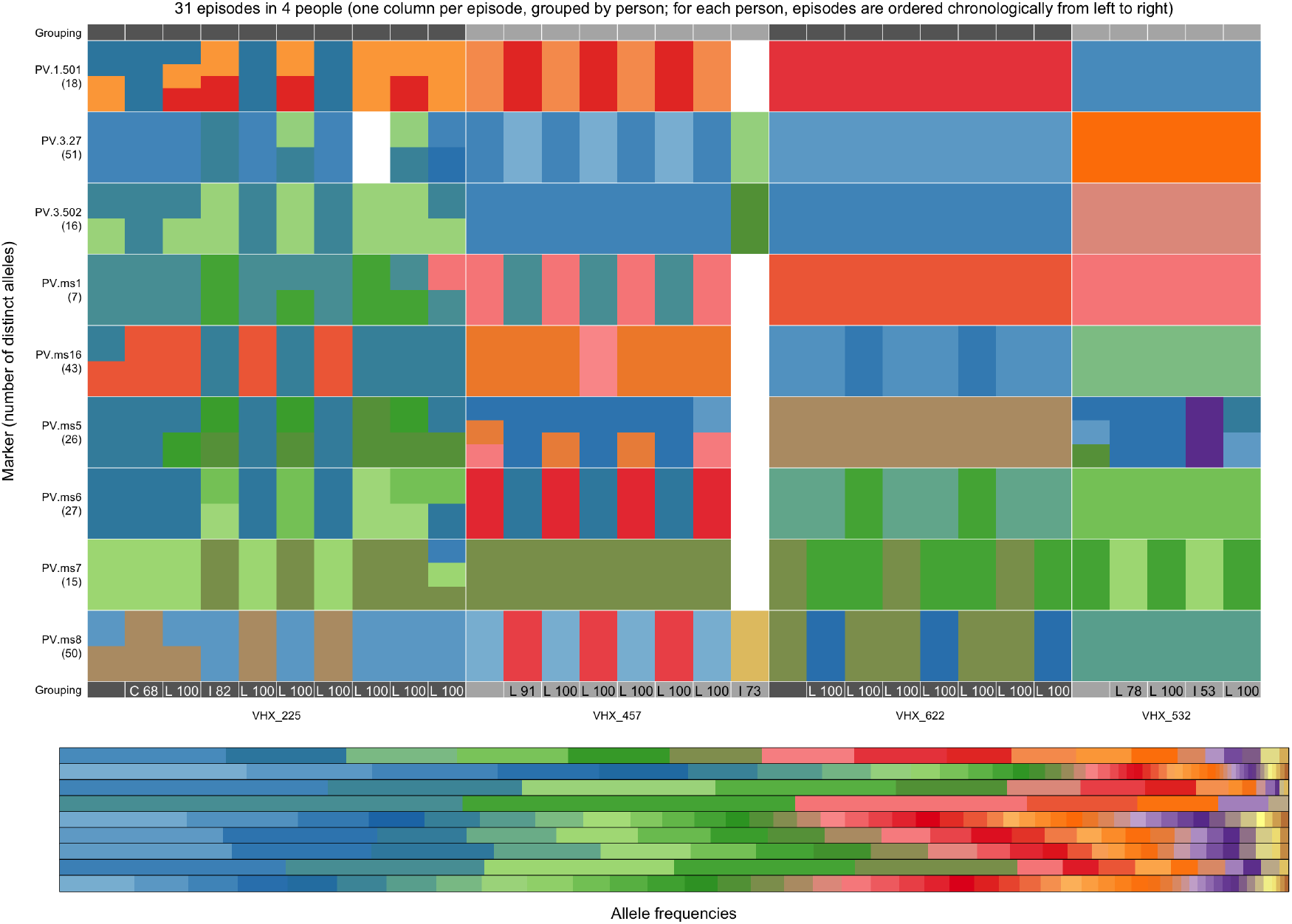
*P. vivax* microsatellite data on selected individuals. This figure was generated using the `plot_data()` function of the Pv3Rs R package. Main plot: alleles (colours), at different markers (rows), detected in different episodes (columns), grouped by person (dark/light alternating grey horizontal ribbon). Per-person episodes are plotted from left to right in chronological order. Where multiple alleles are detected at a single marker within an episode, the corresponding grid element is subdivided vertically into different colours. The legend depicts per-marker alleles (sorted by descending frequency) for each marker in the same vertical order as the main plot, with areas proportional to allele frequencies, so that common alleles (left) occupy larger areas than rarer alleles (right). This plot shows data on 31 episodes in four people. At the bottom of each recurrence, the posterior probability (×100) of the maximum a posteriori state (C, L and I, for recrudescence, relapse and reinfection, respectively) is stated. Probabilities were computed using the Pv3Rs model.

### 2.3 Visual inference

Figure 1 depicts microsatellite data for four example participants from the VHX trial. We can infer visually that these participants have relapsing parasites: highly structured, repetitive patterns suggest recurrent parasites are drawn from per-participant subpopulations. The prototype was built to systematically quantify these patterns using statistical inference. However, reliance on numerics alone is unwise. The Pv3Rs R package includes the function `plot_data()` used to generate Figure 1 and thus supports both statistical and visual inference.

In this manuscript, visual inference is primarily used to identify data consistent with half-sibling parasites (allele matching consistent with sibling parasites drawing from three parental alleles at some markers) and/or genotyping errors (occasional appearance of seemingly spurious alleles). We focus on half-sibling parasites and genotyping errors because the Pv3Rs model is misspecified when fit to data of these types and liable to generate erroneously high posterior reinfection probabilities (Foo et al., 2026). Malfunction depends on the balance of evidence for inter-versus intra-sibling relationships, and thus on the frequencies of the alleles that match within versus across episodes; see Pv3Rs online documentation on half-sibling misspecification using simulated data and theoretical analyses. As such, misspecification does not always lead to erroneously high posterior reinfection probabilities. There are several cases of suspected misspecification in Figure 1, including the 3rd, 4th, and 10th episodes of participant VHX_225. Among these episodes, only the 4th episode appears to have an erroneously high posterior probability of reinfection (> 0.815).

### 2.4 Statistical inference

For a single recurrence, there are three recurrence states: relapse (*L*), recrudescence (*C*), and reinfection (*I*). Posterior recurrence state probabilities computed previously using the prototype and the aforementioned microsatellite data were extracted from the open-source repository zenodo.org/records/3368828 (Taylor et al., 2019). Using the same data, posteriors were re-computed using the Pv3Rs model.

For participants with only two genotyped episodes, per-recurrence posterior probabilities are computed by fitting the prototype/Pv3Rs model to paired-episode data. We refer to this as pairwise inference. For participants with multiple genotyped episodes, per-recurrence posterior probabilities that leverage multi-episode data can be computed either by jointly fitting the prototype/Pv3Rs model to data on all episodes together (joint inference) or by using an approximation of joint inference based on a summary of multiple pairwise inferences. Subsections 2.4.1 and 2.4.2 describe each approach and how they were used in conjunction with the prototype and Pv3Rs. Per-recurrence probabilities whose computation overlooks intervening episodes (e.g., comparing a second recurrence directly to baseline rather than to the first recurrence) are not used here nor in Taylor et al. (2019).

Both the prototype and Pv3Rs require as input allele frequency estimates and prior probabilities. Subsections 2.4.4 and 2.4.5 describe how these inputs differed slightly for the prototype and Pv3Rs.

#### 2.4.1 Joint inference

Both Pv3Rs and the prototype generate posterior probabilities of recurrence state sequences when data on more than two episodes are modelled jointly. For example, for two recurrences, there are nine sequences: *CC, CL, CI, LL, LC, LI, IC, IL, II*. Pre-recurrence probabilities of *C, L*, and *I* are computed by summing over sequence probabilities, i.e., the per-recurrence distribution is the marginal of the joint distribution over sequences. For example, the probability of relapse at the *t*th recurrence in a sequence of recurrences is the sum of the probabilities of sequences with relapse at the *t*th position,

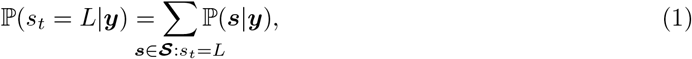

where ***s*** = (*s*_1_, …, *s*_*k*_) is a 1-by-*k* sequence of *k* recurrences, *s*_*t*_ is the recurrence state at the *t*th recurrence, **𝒮** is the collection of all possible sequences of length *k*, and ***y*** = (***y***_*tj*_) is a data matrix of allelic sets observed in *t* = 0, …, *k* episodes across *j* = 1, …, *m* markers. Joint inference is equivalent to pairwise inference when ***y*** is a 2-by-*m* matrix of sets. Otherwise stated, the marginal distribution is equal to the joint, ℙ (***s*** |***y***) = ℙ (*s*_1_| ***y***), because sequences are only one recurrence long.

Computational limits determine how many episodes can be modelled jointly. Using the prototype, joint inference was attempted for per-participant data on two or three episodes with a total multiplicity of infection (MOI) ≤ 6, where MOI is the number of distinct parasite genotypes per infection, equal to the maximum allele count across markers under Pv3Rs, and where total MOI is the sum of MOIs over episodes. However, there were three failed attempts — VHX_239, VHX_52 and VHX_461; see Figure S2. Using the Pv3Rs model, joint inference was attempted if the total MOI ≤ 8 — Pv3Rs enumerates graphs more efficiently than the prototype.

#### 2.4.2 Approximate joint inference

An approximation of joint inference based on a heuristic summary of multiple pairwise inferences can be used to efficiently compute recurrent state probabilities for participants whose data can be partitioned into pairs of episodes with total MOI ≤ aforementioned limits. For all recurrences, probabilities were computed using the Pv3Rs model and the approximate joint inference approach specified by equations (2) to (4):

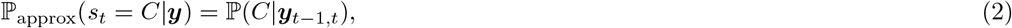

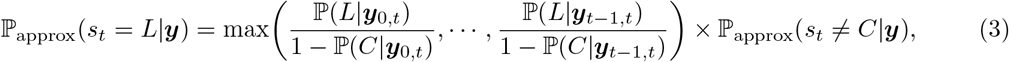

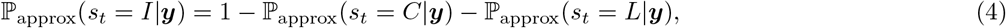

where ℙ (*s*|***y***_*t−*1,*t*_) is the posterior probability of the *t*th recurrence being state *s* based on pairwise inference on data from episodes *t −* 1 and *t*. A similar approach was used with the prototype,

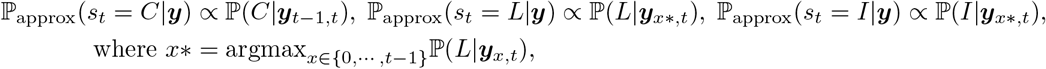

but only for participants where joint inference was not possible (Taylor et al., 2019). Upon normalisation, this approach leads to ℙ_approx_(*s*_*t*_ = *C* |***y***) ≤ ℙ (*C*| ***y***_*t−*1,*t*_). Downwardly weighting recrudescence has little bearing on analyses where a failure is either a recrudescence or a relapse. Nonetheless, this approach is not reproduced here.

#### 2.4.3 Joint versus approximate joint inference

Under a well-specified model, joint inference is superior to approximate joint inference because it simultaneously uses information across all episodes. Equations (2) to (4) synthesise information on episodes 0 to *t* ≤ *k* to infer the state of the *t*th recurrence, but each call to Pv3Rs is pairwise. Moreover, when *t* < *k*, by summing over sequences that extend beyond the *t*th recurrence, joint inference borrows information from episodes that succeed the *t*th. To illustrate how joint inference can allow later episodes to inform earlier ones, consider an extreme scenario: a person with two recurrences where, for all markers typed, a single rare allele in the third episode matches a single rare allele in the first, and where data on the second episode are missing. The probability of these data is low given any sequences that implies the second recurrence is independent of the first episode (*CI, LI, II* and *IC*). Only one sequence among those remaining (*CC, CL, LC, LL*, and *IL*) features reinfection at the first recurrence. As a result, the prior probability of reinfection at the first recurrence exceeds the posterior, despite the first recurrence having no data. Otherwise stated, data on episodes one and three induce a prior-to-posterior decrease in the probability of recurrence at the second episode (first recurrence). Approximate joint inference using the same data returns the prior probability of reinfection for the first recurrence.

#### 2.4.4 Prototype input

In Taylor et al. (2019), uncertainty in allele frequency and time-to-event estimates was propagated through analyses: 100 posteriors were generated per participant by running the prototype with input including 100 different draws from the posterior of an allele frequency model fit to all available data, plus prior probabilities of relapse, recrudescence and reinfection that were either equal (i.e., the uniform prior) or drawn 100 times from the per-recurrence posterior of a time-to-event model; see Taylor et al. (2019). Posteriors given a uniform prior were only generated for recurrences among participants whose data were modelled jointly. In the current study, each set of 100 probabilities per state is summarised by its median and 2.5th and 97.5th percentiles.

#### 2.4.5 Pv3Rs input

Recurrence state probabilities were computed for all recurrences using Pv3Rs with input including the posterior means of the allele frequency model described in Taylor et al. (2019) fit to enrolment data only and either the uniform prior or the time-to-event prior, where relapse, recrudescence and reinfection prior probabilities are equal to mean posterior predictive probabilities computed previously under the time-to-event model described in Taylor et al. (2019). Given there have been no structural changes to the allele frequency and time-to-event models, uncertainty in allele frequency and time-to-event estimates was not re-propagated through Pv3Rs analyses, avoiding unnecessary computation.

### 2.5 Relatedness-based analyses

#### 2.5.1 Classification

In addition to computing recurrence state probabilities using a statistical model, we can assign recurrence state classifications using estimates of genetic relatedness: a recurrence is classified as a relapse and/or recrudescence if its genetic relatedness to any preceding episode within the same participant exceeds that which is expected between episodes that are neither relapse nor recrudescence.

Since relapsing and recrudescent parasites are derived from intra-individual parasite subpopulations, any relapse or recrudescence inferred from data on inter-participant episodes is false (Taylor et al., 2019). A large data set of 250303 inter-participant paired episodes was constructed previously to compute false discovery rates (where a discovery is either a relapse or a recrudescence) using the prototype (Taylor et al., 2019). It contains 247873 pairs with data on at least one marker across both episodes. We used the same data set to determine the expected relatedness between episodes that are certain to be neither relapse nor recrudescence.

First, for each of the 247873 inter-participant episode pairs, we compute inter-episode genetic relatedness using Dcifer, an R package designed to estimate relatedness between malaria parasite infections with MOIs of one or more (Gerlovina et al., 2022). Dcifer generates various measures of inter-episode relatedness, all assuming intra-episode genotypes are unrelated — a simplifying assumption that is almost certainly violated in every setting. Gerlovina et al. (2022) demonstrated through simulation that total relatedness is largely insensitive to model misspecification introduced by intra-episode relatedness. As such, we use total relatedness. Specifically, we use an approximation of total relatedness where all inter-episode genotypes are assumed equal in relatedness. This approximation is quick to compute and generates values that are almost identical to those generated when inter-episode genotypes are allowed to have different levels of relatedness. Second, we base our expected genetic relatedness on some percentile of the distribution of 247873 inter-participant relatedness measures. More specifically, we use the 100(1− *α*) percentile, where *α* = 0.05.

To classify each recurrence, we compute approximate total relatedness between it and all the preceding episodes within the same participant, then take the maximum. For any recurrence whose maximum exceeds the 100(1 − *α*) percentile of the inter-participant distribution, we reject the null hypothesis that it is neither a relapse nor a recrudescence.

#### 2.5.2 Outlier detection and correction

The posterior probability of relapse or recrudescence (i.e., the probability of relapse plus the probability of recrudescence or one minus the probability of reinfection) computed using the Pv3Rs model with a uniform prior was plotted against maximum approximate total relatedness to identify cases where Pv3Rs misspecification likely leads to spuriously high probabilities of reinfection. Posterior probabilities computed using uniform priors are used for outlier detection because the artificial and equal prior facilitates the interpretation of the posterior; there is a clear trend with genetic relatedness, which — like the uniform prior approach — contains no time-to-event information. To see if any outliers require correction, we then plotted posterior probabilities computed using time-to-event priors against genetic relatedness. Among the outliers detected in the first plot, only those that remain outlying after in this second plot are flagged for correction.

Outliers flagged for correction were corrected as follows. Firstly, an interim posterior probability of relapse or recrudescence was assigned by mapping maximum approximate total relatedness onto the trend observed among non-outlying recurrences. Secondly, interim posterior probabilities were adjusted for the time-to-event prior as described in section 2.2 of the supplement of Foo et al. (2026). Corrected posterior probabilities were then used in a sensitivity analysis of radical cure failure rates; see below.

### 2.6 Radical cure failure rate

As in Taylor et al. (2019), the radical cure failure rate was estimated as the average probability of failure in 853 PQ-treated study participants with 677 patient follow-up years (VHX and BPD combined). Since radical cure is designed to clear both liver and blood stage parasites, a failure is defined as either a relapse or a recrudescence. The probability of any failure per participant is one minus the product of per-recurrence reinfection. We note that this assumes independence between recurrences in participants with more than one recurrence. (Aside: we could relax this assumption for participants with more than one recurrence whose data were modelled jointly by computing the probability of any failure as one minus the probability of the all-reinfection recurrence sequence; however, for Pv3Rs-computed probabilities, this makes a total difference of 0.0017 summed across 57 participants.) Reinfection probabilities generated by the time-to-event model are used where genetically informed probabilities are unavailable (e.g., episode 2 of VHX_419) and for censored observations (i.e., participants who did not experience a recurrence insofar as they were followed up). Genetically informed reinfection probabilities are computed using prior probabilities generated by the time-to-event model. Radical cure failure rates were estimated using Pv3Rs-computed probabilities both before and after the outlier correction described above. In Taylor et al. (2019), 95% credible intervals were derived from the propagation of variation in the time-to-event prior and allele frequency input. Here, they are given by the 2.5 and 97.5 percentiles of the Poisson-Binomial distribution over per-trial failure counts, computed using the `qpbinom()` function of the R package PoissonBinomial (Junge, 2026). As such, the 95% credible intervals for prototype-derived rates reported in (Taylor et al., 2019) differ very slightly to those below.

### 2.7 False discovery

#### 2.7.1 False failure classification and probability

Assuming recurrence states are equally likely *a priori*, we computed recurrence state probabilities using Pv3Rs for the 247873 inter-participant episode pairs with data on at least one marker across both episodes. All posteriors given inter-participant paired data are computed using a uniform prior, as the time-to-event concept does not apply to this in silico engineered data set. For comparison with Taylor et al. (2019), where the false discovery rate of 2.5% is a false failure classification rate (the number of recurrences whose probability of relapse plus recrudescence exceed an arbitrary classification threshold of 0.7, normalised by the number of inter-participant pairs with prototype-computable probabilities), we computed a false failure classification rate using the same arbitrary threshold.

In addition, we computed a false failure probability (one minus the probability of reinfection averaged over all inter-participant pairs with computable probabilities) using Pv3Rs, avoiding reliance on an arbitrary classification threshold. Unless the prior on relapse is zero, the false failure probability will never be zero because both relapse and reinfection are viable when data are compatible with reinfection. Otherwise stated, given a non-zero prior probability of relapse, Pv3Rs returns a non-zero posterior probability of relapse when data are consistent with reinfection. MOIs induce upper bounds on posterior probabilities of reinfection when data are consistent with reinfection (see online article Understand graph-prior ramifications for more details). As such, we can compute the irreducible false failure probability (the probability we would obtain with theoretical data that are optimally informative) using Pv3Rs-determined MOIs of inter-participant paired episodes, and compare it to the probability based on empirical data. Specifically, the irreducible false failure probability is one minus the upper bound on the posterior probability of reinfection averaged over all inter-participant pairs with computable probabilities.

#### 2.7.2 False non-failure probability

To estimate the false non-failure probability (i.e., the probability of missing a true relapse or recrudescence) in endemic residents, we need a study design that removes the risk of reinfection; see Popovici et al. (2018, 2026). Instead, we approximate the false non-failure probability assuming recurrences classified as relapse or recrudescence using genetic relatedness are true failures. More specifically, we estimate the false non-failure probability by summing the posterior probability of reinfection over the recurrences classified as relapse or recrudescence using genetic relatedness. We do this for posteriors computed using both the uniform and the time-to-event prior.

## 3 Results

### 3.1 Computational capability

The Pv3Rs model outperforms the prototype regarding both the number of study participants whose data were modelled jointly and the number of recurrences whose recurrent state probabilities were computed using either a joint or approximate joint approach; see Table 2 and Figure S2.

**Table 2:**
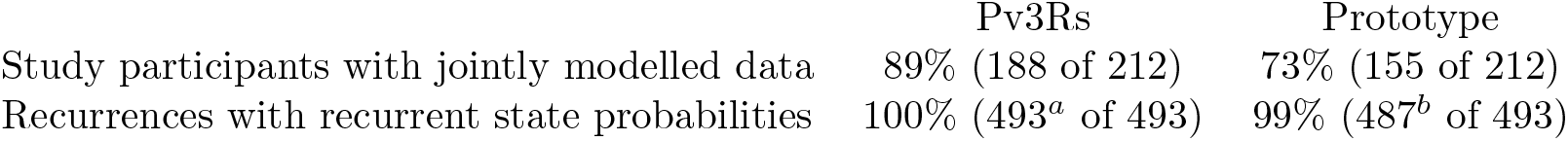
Computational capability. ^*a*^Includes 313 recurrences among 188 participants with jointly-modelled data. ^*b*^Includes 183 recurrences among 155 participants with jointly-modelled data.

### 3.2 Probabilities of relapse, recrudescence and reinfection

#### 3.2.1 Probabilities computed using both Pv3Rs and the prototype

Overall, probabilities computed using the Pv3Rs model are similar to those computed using the prototype (Figure S3). They concentrate on either relapse, relapse and recrudescence, or relapse and reinfection. Few recurrences in PQ-treated participants have high recrudescence or relapse probabilities, especially when time-to-event information is used a priori. Many more recurrences in PQ-untreated participants have high recrudescence or relapse probabilities, especially when time-to-event information is used a priori.

Despite overall similarity, probabilities differ on a recurrence-to-recurrence level. Given the uniform prior, the posterior probability of reinfection is almost always elevated by the Pv3Rs model (Figure S4). Seven of 183 (3.8%) recurrences have posterior probabilities that differ by more than 0.25 given the uniform prior. Four of 487 (<1%) recurrences have posterior probabilities that differ by more than 0.25 given the time-to-event prior. These four include two of the seven recurrences whose posterior probabilities differ by more than 0.25 using the uniform prior.

Visual inference suggests that among the nine recurrences with discrepant probabilities, Pv3Rs relapse probabilities are unreasonably low in two cases: VHX_56_2 (PQ-treated) and VHX_91_2 (PQ-untreated). In both cases, recurrence appears compatible with half sibling parasites: allele matching is high and consistent with recurrent parasites drawing from three parental alleles at some markers (Figure S5).

#### 3.2.2 Probabilities computable using Pv3Rs only

Data from six participants with a recurrence whose probabilities could not be computed using the prototype are shown in Figure S6. When recurrence states are considered equally likely a priori, Pv3Rs estimates with more than 80% probability that four of the six recurrences are either relapse or recrudescence. VHX_39_2 (PQ-untreated) is estimated to be a relapse with 51% probability and a reinfection with 49% probability and appears to be a possible case of half-sibling misspecification. All recurrences are estimated to be relapse with more than 99% probability using the time-to-event prior.

#### 3.2.3 Joint versus approximate joint inference with Pv3Rs

There were 182 recurrences in 57 study participants with genetic data on more than one recurrence (i.e., where joint inference is not pairwise) that could be modelled jointly using Pv3Rs and thus compared non-trivially to approximate joint inference.

Given the uniform prior, a slight elevation in recrudescence probability is observed under joint inference. This is consistent with the Pv3Rs graph prior: as the number of episodes (and hence the graph space) expands, the fraction of graphs compatible with recrudescence decreases, so that conditional on observing clonal matching, the weight of evidence for recrudescence strengthens; again, see the online article Understand graph-prior ramifications. This does not necessarily mean more biological evidence of recrudescence; it is a structural property of the model’s state space.

Given either prior, joint inference also elevates posterior reinfection probability. Elevated reinfection probabilities are a logical outcome of the additional conditioning on all available earlier data that joint inference provides. When data on multiple episodes are modelled jointly, each unmatched episode provides extra evidence against relapse or recrudescence — a greater number of opportunities to match strengthens inference of independence, i.e., reinfection. In addition, when the model is fit to data on multiple episodes, the model is more likely to encounter alleles that break the assumption that siblings draw their alleles from at most two parental gametes.

Six posterior probabilities computed using a uniform prior differ by more than 0.25 (left column, Figure S7). Visual inference suggests all of these cases are likely relapses with half sibling parasites (high allele matching consistent with recurrent parasites drawing from three or more parental alleles) (Figure S8). In five of these six cases, approximate joint inference generated a higher posterior relapse probability than joint inference: approximate joint inference is less sensitive to half-sibling misspecification if a study participant’s data can be paired in a way that strengthens evidence of inter-versus intra-sibling relationships. When posteriors were computed using the time-to-event prior, only one probability, that of VHX_419_6, differed by more than 0.25 (right column, Figure S7). In the case of VHX_419_6, the prior relapse probability of 0.97 has a stronger impact on the posterior computed using the approximate joint approach.

#### 3.2.4 Corroboration using genetic relatedness

In addition to computing posterior probabilities of recurrence states, we can classify recurrences as either reinfection or not using a measure of genetic relatedness. More precisely, for a given level of significance (e.g., *α* = 0.05), we can accept or reject the null hypothesis that a recurrence is neither a relapse nor a recrudescence by comparing its measure of genetic relatedness to a threshold, where the threshold is the 100(1 *− α*) percentile of the distribution of proximities between all inter-participant episode pairs (pairs where the second episode is neither a relapse nor a recrudescence). Figure 2 shows the probability of relapse or recrudescence against maximum approximate total relatedness; see methods. We highlight two important observations:

**Figure 2.**
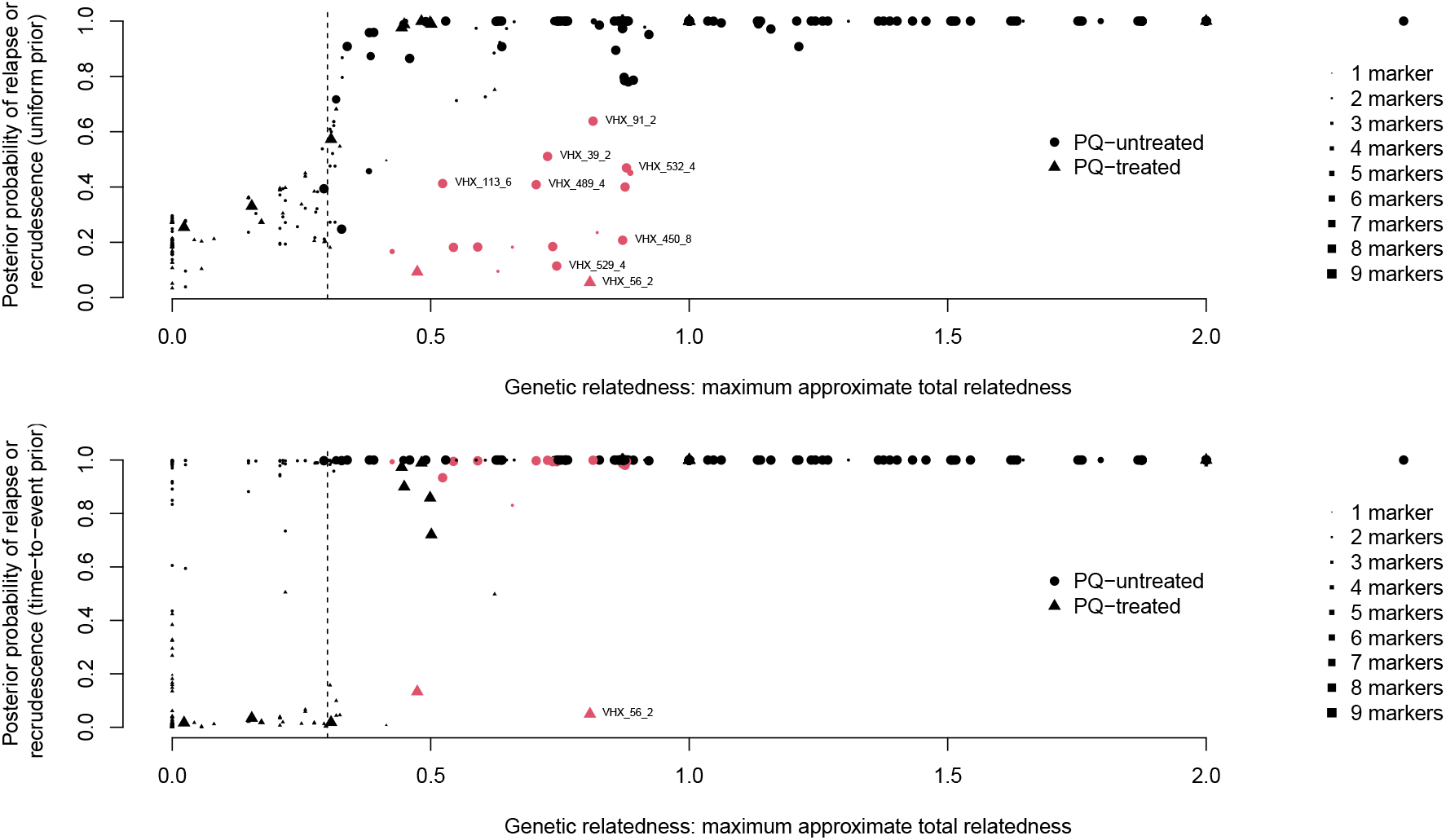
Probability of recrudescence plus relapse against genetic relatedness. The dashed vertical line marks the threshold used to reject the null hypothesis that a recurrence is neither a recrudescence nor a relapse with 5% probability of rejecting the null when it is actually true. Red points appear not to follow the sigmoid trend between relatedness and probability computed using a uniform prior. Labels (study acronym underscore participant ID underscore episode number) locate recurrences where misspecification has been suspected already: VHX_56_2 and VHX_91_2 whose relapse probabilities computed using the Pv3Rs model were more than 0.25 lower than those computed using the prototype; VHX_39_2 among recurrences whose probabilities were not computable using the prototype; and VHX_113_6, VHX_450_8, VHX 489 4, VHX_529_4, and VHX_532_4 among the recurrences whose relapse probabilities where more than 0.25 lower for joint versus approximate joint inference using Pv3Rs. Among these cases of suspected misspecification, only VHX_56_2 has highly discrepant probabilities when probabilities are computed using the time-to-event prior (bottom plot).

1. Among recurrences with low relatedness (those on the left of the classification threshold), the average posterior probability of relapse is not zero. This is because the Pv3Rs model recognises that relapsing parasites may be genetically unrelated to their predecessors (both reinfection and relapse are viable states when genetic data are compatible with reinfection), whereas classification using genetic relatedness does not: every instance of low relatedness is classified categorically as reinfection.
2. We recover a sigmoid relationship between per-recurrence posterior probabilities computed using the uniform prior and genetic relatedness. The inflection point approximately aligns with the relatedness-based classification threshold whose significance level is 5%. Various outliers with high relatedness and low posterior probability do not follow this trend; they are highlighted in red in Figure 2. Among these outliers, we find all the aforementioned cases where misspecification likely results in malfunction of the Pv3Rs model; they are labelled in Figure 2. Visual inferences suggests there is a strong case for misspecification among most of the outliers that are not already mentioned (Figure S9). Only two of the outliers remain outlying when time-to-event information is accounted for *a priori*; see bottom plot, Figure 2. One, VHX_56_2, is among the aforementioned cases of suspected malfunction due to misspecification (see Figure S5); the other, BPD_577_2, was not already mentioned and thus features in Figure S9. Both VHX_56_2 and BPD_577_2 are in PQ-treated participants and thus their correction (or not) has a potential bearing on the radical cure rate.

### 3.3 Radical cure failure rate

As in Taylor et al. (2019), radical cure failure rates are estimated by summing over posterior probabilities of failure in PQ-treated participants. Given the aforementioned outliers, VHX_56_2 and BPD_577_2, failure rates based on probabilities generated by the Pv3Rs model were computed before and after outlier correction: following the sigmoid trend by eye, interim posterior probabilities of 0.990 relapse plus recrudescence (i.e., 0.010 reinfection) were attributed to outliers BPD_577_2 and VHX_56_2 (Figure S10). After adjusting for time-to-event priors, which were 0.400 reinfection for BPD_577_2 and 0.527 reinfection for VHX_56_2, corrected posterior probabilities of 0.007 reinfection for BPD_577_2 and 0.011 reinfection for VHX_56_2 were obtained.

Despite some discrepant probabilities of failure at the per-participant level (Figure S11), per-study failure rates estimated using Pv3Rs-computed probabilities are similar to those estimated using prototype-computed probabilities (Table 3). Before the correction of the two outliers, the large and reasonable Pv3Rs-computed failure rate for VHX participant 583 (VHX_583 has a recurrence whose probability was not computable using the prototype) compensates for the unreasonably low Pv3Rs-computed failure rate for VHX participant 56 (one of the two outliers); two BPD participants (both of whom seem to have reasonable recurrence state probabilities under Pv3Rs; see Figures S4 and S5), also counterbalance one-another. After correction of the two major outliers, the unreasonably low failure rate for VHX participant 56 disappears and a high and reasonable failure rate for participant BPD_577 is added, elevating the failure rate, but only slightly.

**Table 3:** Mean reinfection-adjusted failure rates of high-dose primaquine (Poisson-Binomial 95% credible interval) computed using recurrence state posterior probabilities generated by the prototype versus Pv3Rs. Probabilities generated by Pv3Rs were computed before and after correcting two major outliers, BPD_577_2 and VHX_56_2.

|  | BPD | VHX | BPD and VHX combined |
| --- | --- | --- | --- |
| Prototype | 3.05 (2.29, 3.82) | 2.37 (1.01, 4.04) | 2.89 (2.23, 3.63) |
| Pv3Rs (outliers uncorrected) | 3.02 (2.29, 3.82) | 2.33 (1.01, 4.04) | 2.86 (2.23, 3.52) |
| Pv3Rs (outliers corrected) | 3.15 (2.44, 3.97) | 2.81 (1.52, 4.55) | 3.07 (2.46, 3.75) |

### 3.4 False discovery rates

#### Prototype false failure classification

Previously, a false failure classification rate of 2.5% was reported, where a recurrence was classified as a failure if its posterior probability of relapse plus recrudescence (computed using a uniform prior) exceeded 0.7 (an arbitrary threshold). This rate was averaged over 249540 inter-participant episode pairs for which a posterior was calculable using the prototype. These 249540 pairs include 1667 pairs without data on a common marker across episodes. Pairs without data on a common inter-episode marker are liable to return a posterior close to the prior (see [Data are incomparable across episode](https://aimeertaylor.github.io/Pv3Rs/articles/posterior-probabilities.html#incomparable) in the article [Understand posterior probabilities](https://aimeertaylor.github.io/Pv3Rs/articles/posterior-probabilities.html) on Pv3Rs GitHub pages) and thus be classified as a failure. Re-computation of the prototype failure rate using the 247873 pairs with data on one or more common markers generates a false failure classification rate of 2.4%.

#### Pv3Rs false failure classification and probability

With the the same arbitrary threshold, the false failure classification rate using Pv3Rs-computed probabilities is 1.8% averaged across all 247873 inter-participant episode pairs with data on one or more common markers across episodes. Using Pv3Rs-computed probabilities, we also computed a false failure probability of 25.3%, avoiding reliance on an arbitrary threshold of 0.7. It exceeds the irreducible failure probability of 20.4% by only 4.9%, and is very close to the average 25.0% probability of relapse or recrudescence computed using the uniform prior among recurrences classified as reinfection using genetic relatedness.

#### Pv3Rs false non-failure probability

Despite numerous outliers (Figure 2), none of which were corrected in the calculations of the following rates, the average probability of reinfection is low among recurrences classified (using genetic relatedness) as relapse or recrudescence: 0.08 when probabilities are computed using a uniform prior and 0.03 when probabilities are computed using a time-to-event prior.

## 4 Discussion

We revisit *P. vivax* molecular correction for the VHX and BPD trials. Despite some differences at the per-recurrence and thus per-participant level, study-level reinfection-adjusted failure rates of high-dose primaquine are nearly identical to those reported previously (Taylor et al., 2019).

We used Pv3Rs for both statistical (model-based) and visual inference of recurrence states and Dcifer for relatedness-based analyses (Foo et al., 2026; Gerlovina et al., 2022). These state-of-the-art methods are both available as R packages distributed on CRAN (Taylor and Foo, 2025; Gerlovina, 2025). Dcifer is a general purpose tool, designed for estimating relatedness between malaria parasite infections, including those that are polyclonal. The Pv3Rs model was designed specifically for *P. vivax* molecular correction. That said, the data plotting function could be used to visualise data on any malaria parasite infection, including those generated for *P. falciparum* molecular correction. It is an extension of the prototype that was built to analyse the VHX and BPD microsatellite data, which feature alleles detected at ≤ 9 markers (Taylor et al., 2019). To the best of our knowledge, all other models designed to infer per-participant *P. vivax* recurrence states are limited to data on a single marker (White et al., 2018; Lin et al., 2020; Liu et al., 2021; Jiang et al., 2022).

The Pv3Rs model is computationally more capable than the prototype, computing recurrence state probabilities for more recurrences and with joint inference for more participants. Nevertheless, the Pv3Rs model remains limited to data on episodes whose MOIs sum to ≤ 8, as the model’s graph space grows rapidly with increasing MOI. Probabilities are broadly consistent with those computed previously. A small but systematic elevation in reinfection probability aligns with the correction of a mathematically convenient but biologically meaningless assumption built into the prototype — an assumption expected to disproportionally bias reinfection probabilities downwards. The correction of this assumption in the Pv3Rs model introduced a new assumption: that all sibling parasites draw their alleles from at most two parental gametes. This assumption renders Pv3Rs more susceptible to biological and technical misspecifications, a problem illustrated by several outliers. Limited scalability, sensitivity to misspecification, and other statistical constraints (see Table S1) merit future work to improve Pv3Rs. Likewise, advances in our understanding of relapse merit future work. For example, the Pv3Rs model assumes time-to-event and genetic data are independent; a recent study challenges that assumption, with faster parasite growth rates measured among relapsing parasites that are unrelated versus related to those detected previously (Popovici et al., 2026).

Another departure from biological realism, present in both the Pv3Rs model and the prototype, is the assumption that all graphs compatible with a given recurrence state are *a priori* equally likely. In reality, homologous relapses predominate early in life; while heterologous ones accumulate with exposure (Imwong et al., 2007, 2012). Under a uniform prior on recurrence states, this assumption generates posterior upper bounds on the probability of reinfection and recrudescence that capture the geometry of the model’s internal graph space. Incorporating time-to-event information shifts these bounds towards more biologically meaningful ones, but does not capture infection history: an infant and an adult who both have informative data compatible with reinfection, the same time-to-event prior, and identical MOIs across episodes will obtain the same residual posterior probability of relapse, even though epidemiologically the uncertainty between reinfection and relapse would be greater in the adult. Incorporating exposure-dependent structure into the prior on graphs would bring the model closer to biological plausibility but risks increasing model complexity and thus error.

In this study, we focus on ways to optimise the utility of the current version 1.0.0 of Pv3Rs (DOI: 10.32614/CRAN.package.Pv3Rs). Estimates of genetic relatedness proved valuable for identifying outliers, enabling us to harness the advantages of the Pv3Rs model (its ability to model relapsing parasites that are genetically unrelated to their predecessors, to output probabilistic rather than categorical results, and to incorporate prior information such as that derived from time-to-event data) despite its sensitivity to misspecification. More specifically, comparing relatedness to per-recurrence posterior probabilities computed using uniform priors revealed a sigmoid trend, enabling the detection outliers, as well as the correction of outliers that remain outlying when the posterior is computed using time-to-event priors. Interim posterior probabilities corrected under the uniform prior assumption were then transformed using time-to-event priors. This transformation worked because joint inference was pair-wise for the outliers detected (if marginal and joint distributions differ, one can correct the marginal, but not reconstruct the joint, which is required for transformation; see section 2.2 of the supplement of Foo et al. (2026)). When an outlier is detected in a participant with more than two genotyped episodes, approximate joint inference alone (i.e., without correction) can sometimes lead to more logical results, e.g., the 8th episode of VHX 450, Figure S8. Otherwise, correction of one or more pairwise inferences could be combined with approximate joint inference.

Together, Pv3Rs and Dcifer have both been used twice before for *P. vivax* recurrence state inference. Rosado et al. (2026) used both to demonstrate the utility of a newly developed amplicon deep-sequencing assay, PvAmpSeq, showing that integrated analysis of relatedness and recurrence state probabilities enables broadly consistent identification of relapse and reinfection patterns across two different transmission settings, the Solomon Islands and Peru. An inter-participant paired episode data set was used to compute the probability of false discovery (recrudescence or relapse): in the Solomon Islands, the false discovery probability exceeded an irreducible probability of 15.7% by 4.9 (same excess reported here); in Peru, it exceeded an irreducible probability of 16.2% by 10.8. Tadesse et al. (2026) combined data generated using PvAmpSeq with Dcifer and Pv3Rs to analyse Ethiopian *P. vivax* recurrences, showing that about 46% of genotyped recurrent infections were relapses, and that relapse infections produced fewer infected mosquitoes than reinfections after adjustment for gametocyte density. These findings link genetic recurrence classifications to transmission potential and highlight the need for effective radical-cure strategies to limit transmission.

To conclude, this is not the first study featuring both Pv3Rs and Dcifer. However, it is the first to demonstrate with real data how the Pv3Rs model is susceptible to misspecification, and how relatedness estimated using Dcifer can be used to both detect and correct problematic cases of misspecification. It is also the first study where inter-participant relatedness estimates are used to accept or reject the null hypothesis that a *P. vivax* recurrence is neither a relapse nor a recrudescence, inspired a similar approach used for *P. falciparum* classification (Plucinski and Barratt, 2021). As such, we hope that this study will help future studies optimise their use of both Pv3rs and Dcifer.

## 5 Data and code availability

All *P. vivax* microsatellite data are included with the Pv3Rs R package (version 1.0.0; CRAN DOI 10.32614/CRAN.package.Pv3Rs). All other input data, analysis scripts, and code for reproducing the results and figures are available at github.com/aimeertaylor/PvMolCorrection/releases/tag/v0.0.0.90.

## Supplement

Unless otherwise stated, all per-participant genetic data plots were generated using `Pv3Rs::plot_data`; see Figure 1 for a general description.

**Figure S1.**
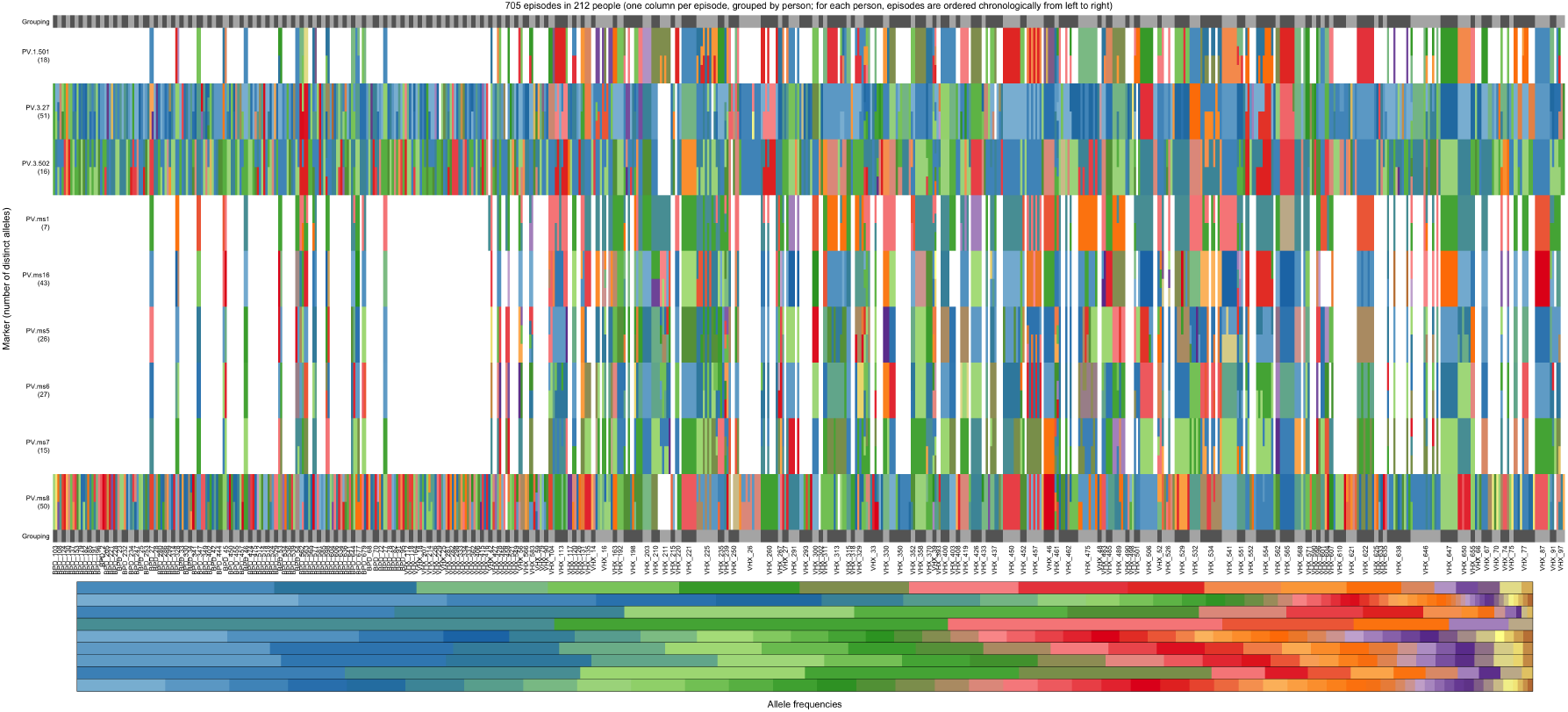
All available microsatellite data on VHX and BPD trial participants with two or more episodes genotyped. Participant IDs are hard to read but included. Data on samples from PQ-treated participants are on the left hand side (pre VHX_104, including all BPD participants). Data on samples from PQ-untreated participants are on the right hand side (VHX_104 onwards).

**Figure S2.**
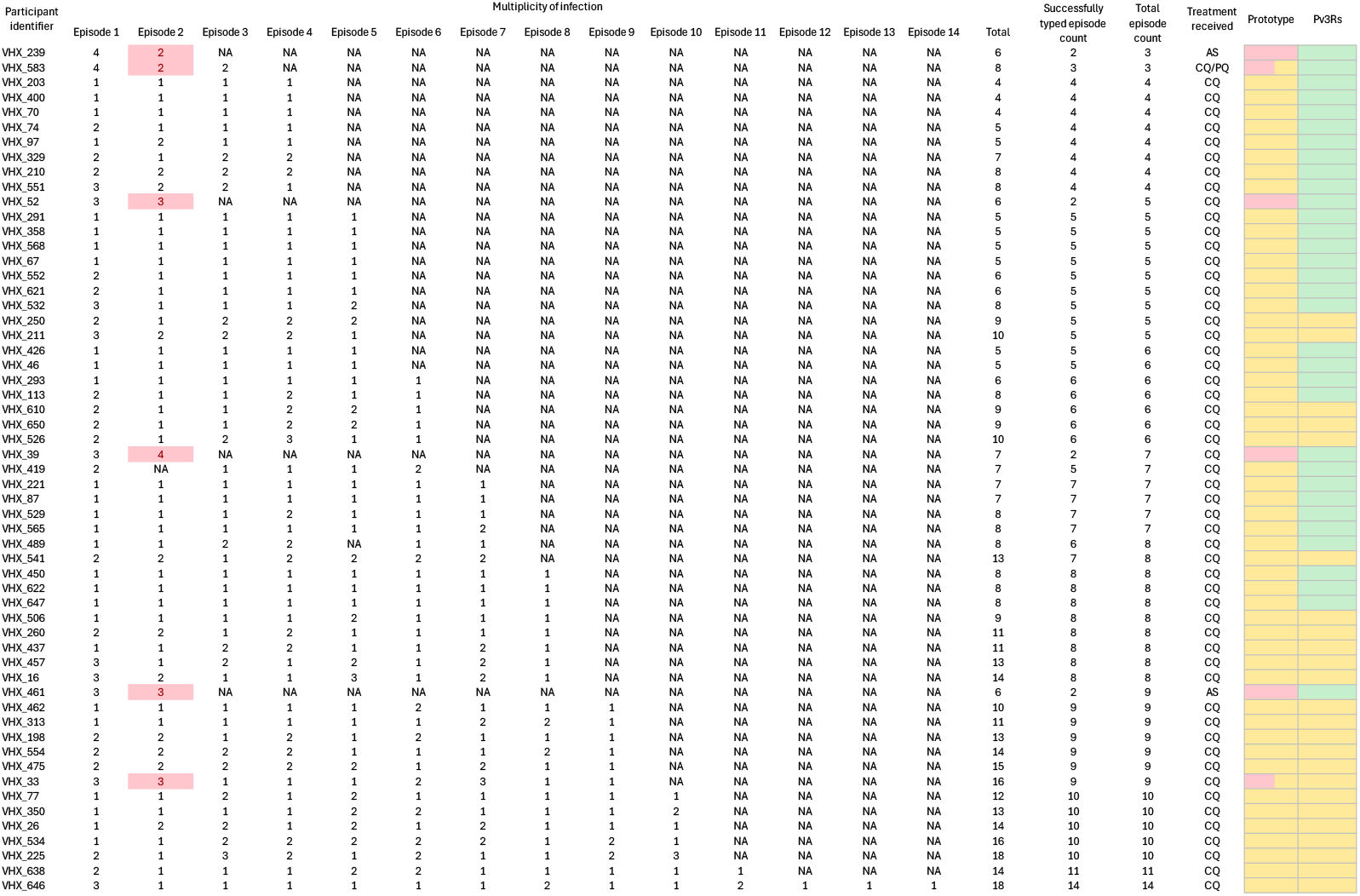
A visual summary of Pv3R’s enhanced computational capability, focusing on 57 VHX trial participants whose data were not modelled jointly under the prototype. Episodes that were not successfully genotyped have not available (NA) multiplicities of infection. Red marks recurrences (all of which happen to be first recurrences — column entitled Episode 2) and participants (column entitled Prototype) whose data were too complex to model using the prototype. Yellow marks participants whose data were modelled using an approximation of joint inference with either the prototype or Pv3Rs. Green marks participants with jointly modelled data under Pv3Rs. Acronyms: artesunate, AS; chloroquine, CQ; primaquine, PQ.

**Figure S3.**
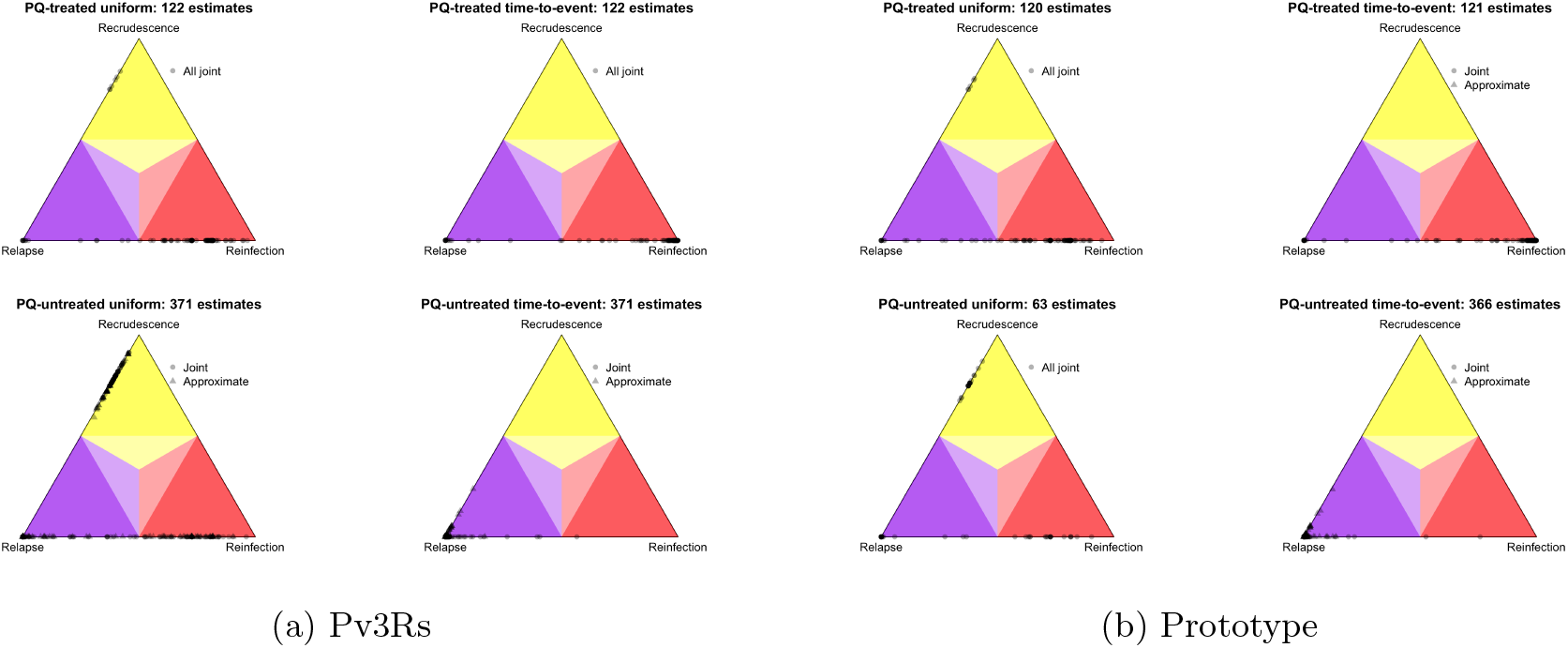
Per-recurrence state probabilities computed using the Pv3Rs model versus the prototype for recurrences in PQ-treated versus PQ-untreated participants assuming either recurrence states are equally likely *a priori* (uniform) or using time-to-event information *a priori* (time-to-event). Joint inference was used to compute probabilities where possible; otherwise, an approximation of joint inference was used. The approximation used with Pv3Rs is not identical to that used previously with the prototype; see methods. Also, although both Pv3Rs and the prototype were run using a uniform and time-to-event prior, specific details differ; see methods. Using the prototype, probabilities assuming uniform recurrent states *a priori* were only generated for 183 recurrences in 155 study participants with data on three or fewer episodes.

**Table S1:** Summary of statistical and computational limitations of the Pv3Rs model.

| Issue | Why it is a limitation | Consequence for inference |
| --- | --- | --- |
| Uniform priors on graph and partition spaces. | Assumes all relationship structures are equally probable, ignoring epidemiological and biological constraints. | Posterior probabilities capture combinatorial volume rather than realistic parasite biology. |
| No observation model. | Ignores genotyping errors. | Produces overconfident posteriors that can misclassify recrudescence as relapse or relapse as reinfection. |
| Fixed allele frequencies. | Treats frequency input as certain. | Ignores uncertainty. |
| Mutual exclusivity of recurrence states. | Disallows mixed-state, concurrent and persistent processes. | Ill-defined posterior when multiple processes contribute to recurrence. |
| Fixed multiplicities of infection. | Conditions on observed allele counts rather than modelling infection process. | Puts hard constraints on graph space and ignores uncertainty. |
| Reliance on exhaustive enumeration of graphs and partitions. | Exact likelihood evaluation requires summing over all compatible graph and partition spaces. | Creates severe computational bottlenecks and limits scalability to data where MOIs are modest. |

**Figure S4.**
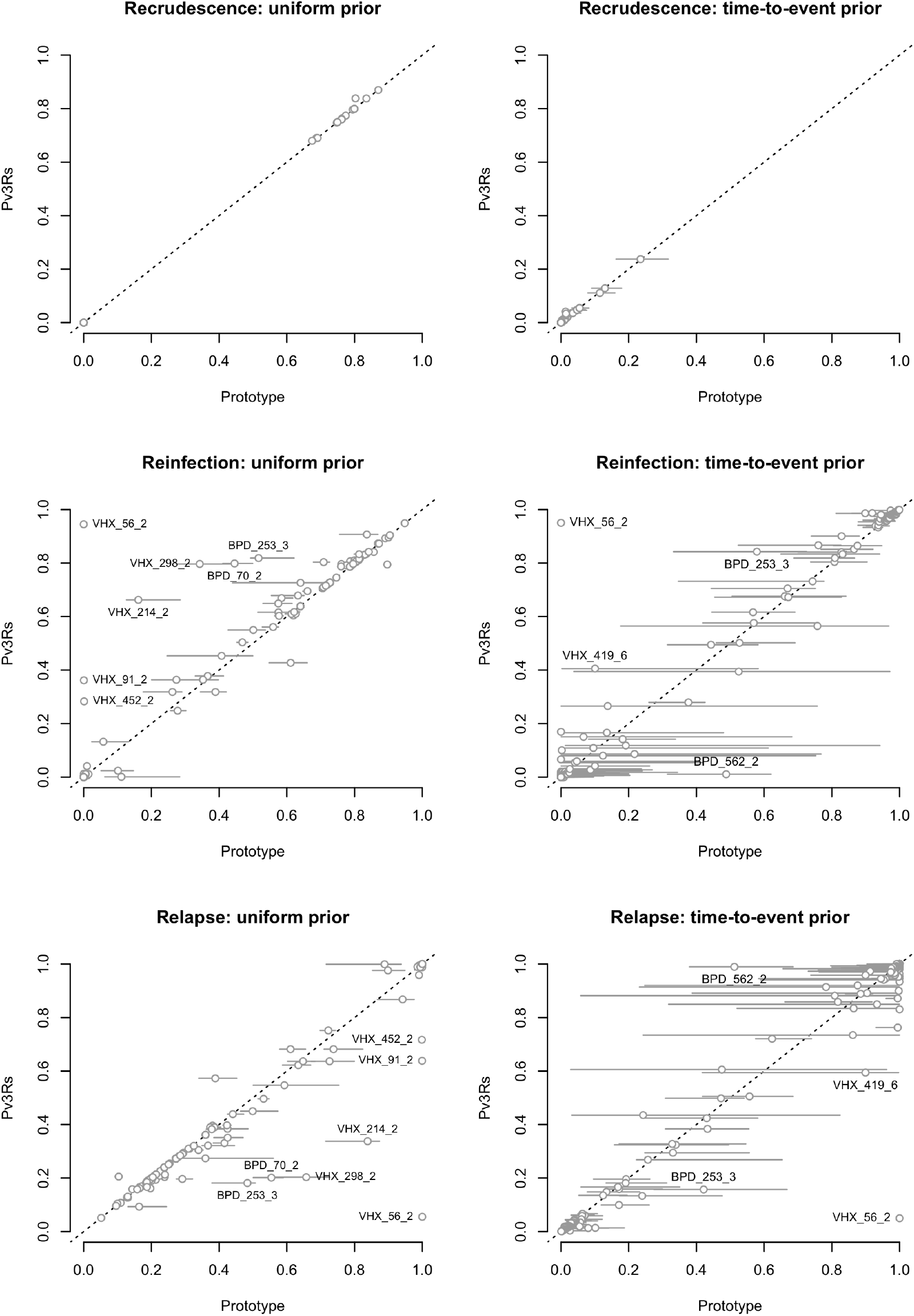
Probabilities computed using both Pv3Rs and the prototype. Pv3Rs probabilities are plotted against the prototype median (grey open circle) with horizontal bars stretching from the 2.5th to 97.5th percentile. Recurrences for which the Pv3Rs probability differs to the prototype median by more than 0.25 are labelled.

**Figure S5.**
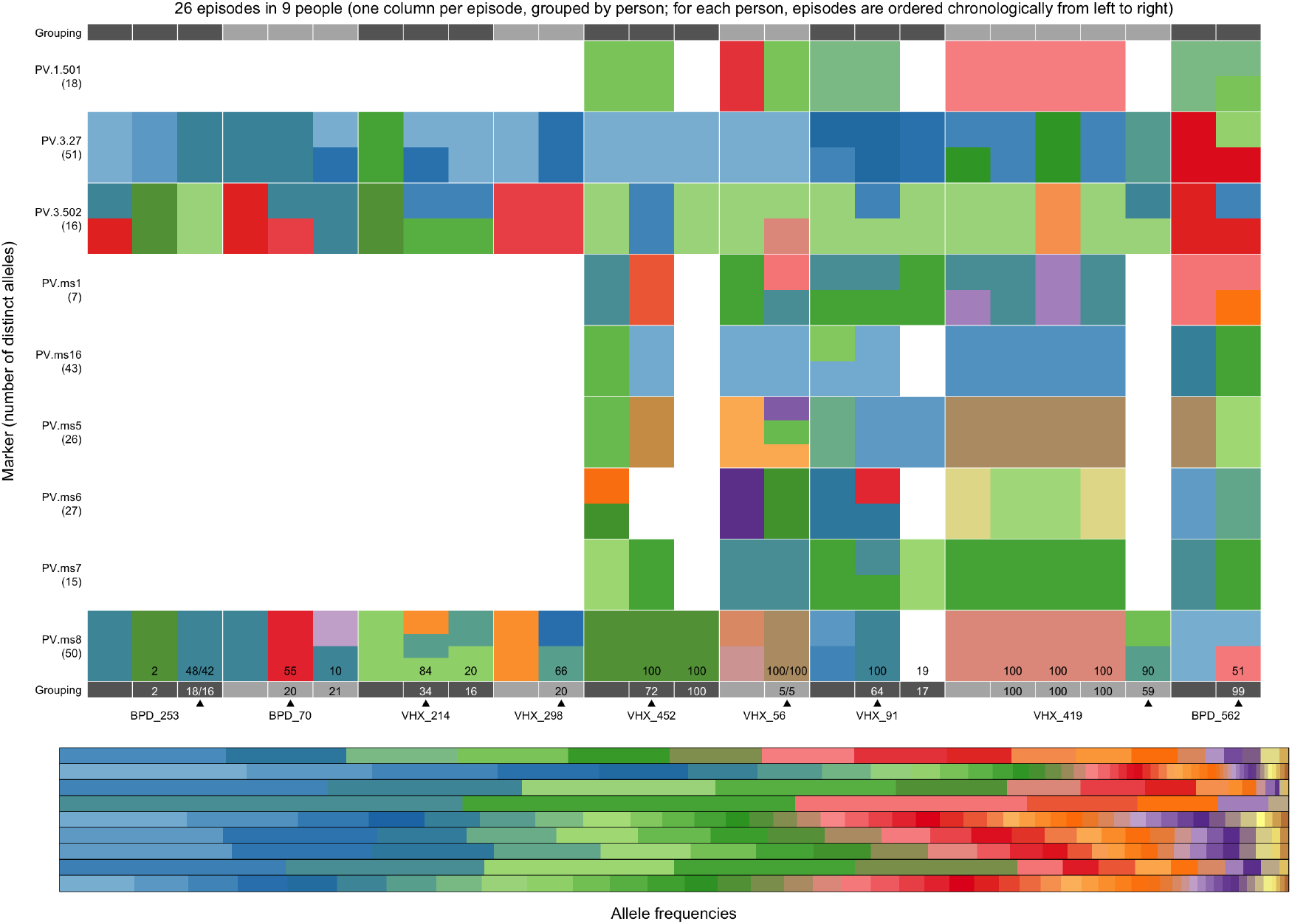
Data from study participants with per-recurrence probabilities computed using the Pv3Rs model and the prototype that differ by more then 0.25. Recurrences whose probabilities differ by more than 0.25 are indicated by an arrow. The Pv3Rs-computed relapse probability (×100) is stated below that computed using the prototype. Stated probabilities were computed using a uniform prior with the following two exceptions: VHX_419, for whom a posterior given a uniform prior was not computed using prototype (using the prototype, posteriors given a uniform prior were only computed where joint inference was possible, which was not the case for VHX_419; see Section 2.4.4 and Figure S2); BPD 562 whose posterior differs significantly only when the time-to-event prior is used (both Pv3Rs and the prototype consider the recurrence of BPD_562 to be a relapse with more than 0.88 probability — prototype — and more than 0.99 probability — Pv3Rs — when states are equally likely a priori; when prior time-to-event probabilities are used, the posterior of the prototype remains closer to the prior, which is more than 92% reinfection). Where probabilities computed using either prior differ by more than 0.25, both probabilities are stated in the following format: posterior given uniform prior ×100 / posterior given time-to-event prior ×100.

**Figure S6.**
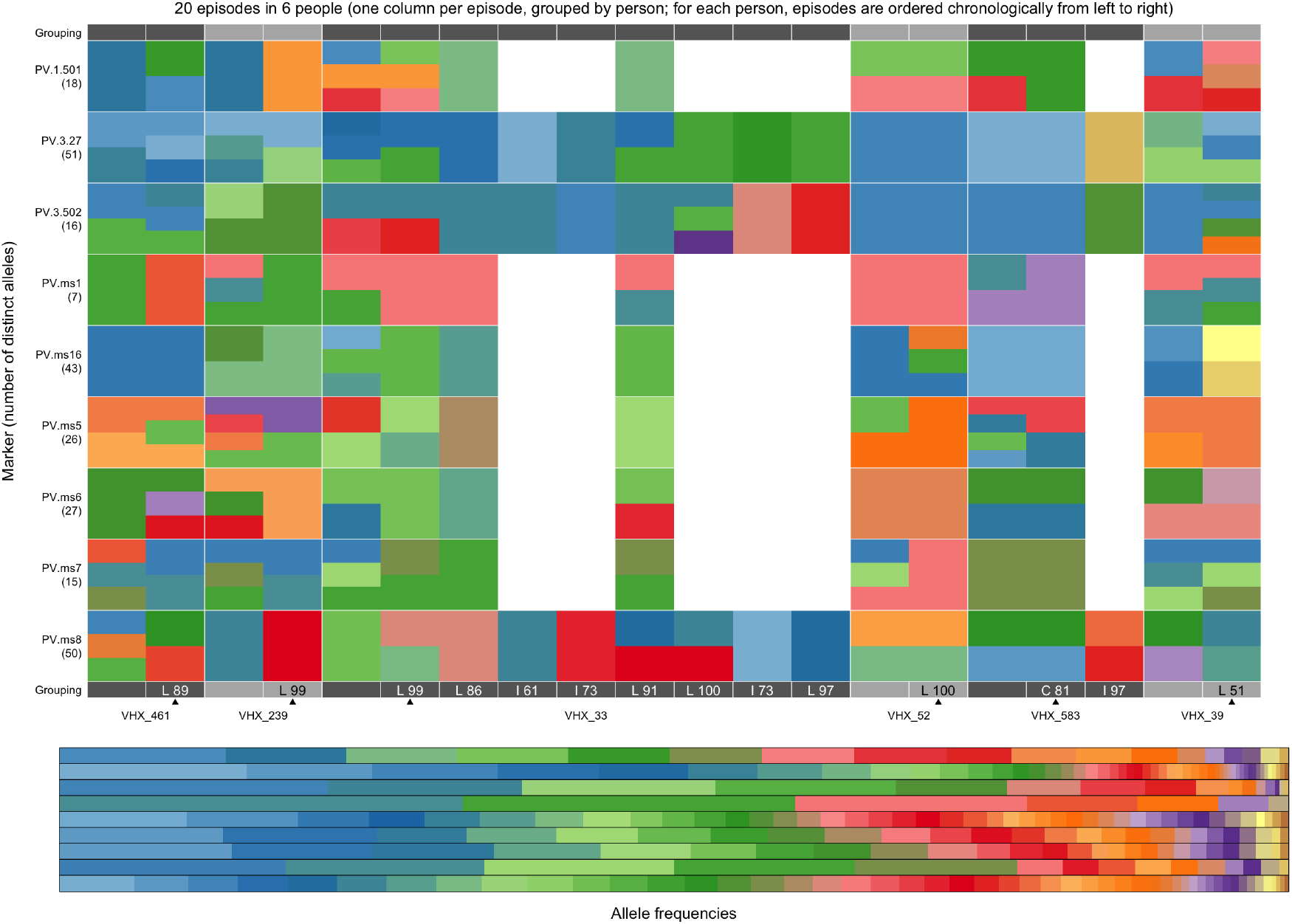
Parasite genetic data from episodes in study participants with a recurrence (indicated by an arrow) whose recurrent state probabilities could not be estimated using the prototype. For each recurrence, the posterior probability (×100) of the most probable state computed using the Pv3Rs model with a uniform prior on recurrent states is stated, where L, C and I are shorthand for relapse, recrudescence and reinfection, respectively.

**Figure S7.**
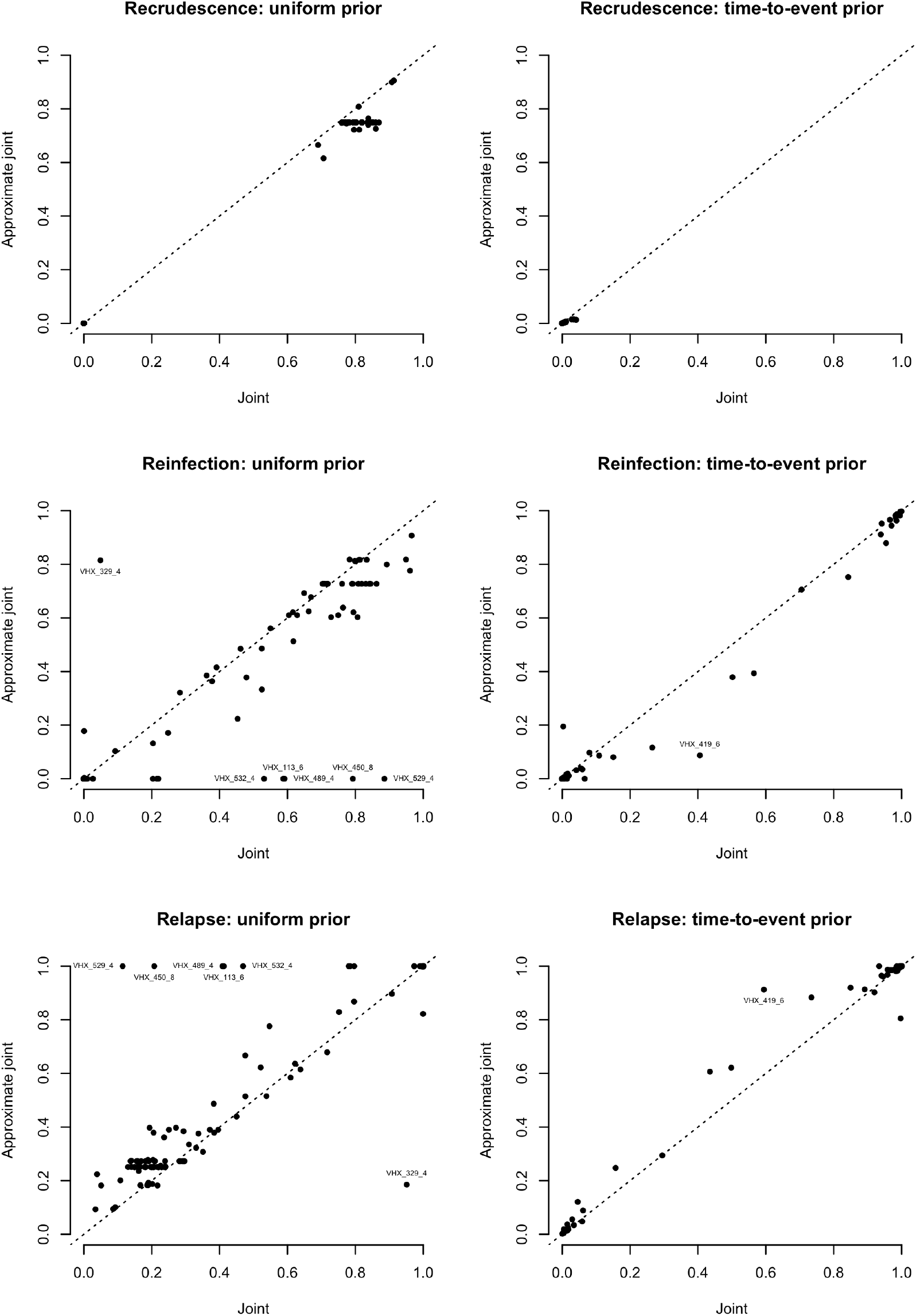
Posterior probabilities for 182 recurrences in 57 study participants with more than one recurrence (i.e., where joint inference is not pairwise) computed using joint versus approximate joint inference with Pv3Rs.

**Figure S8.**
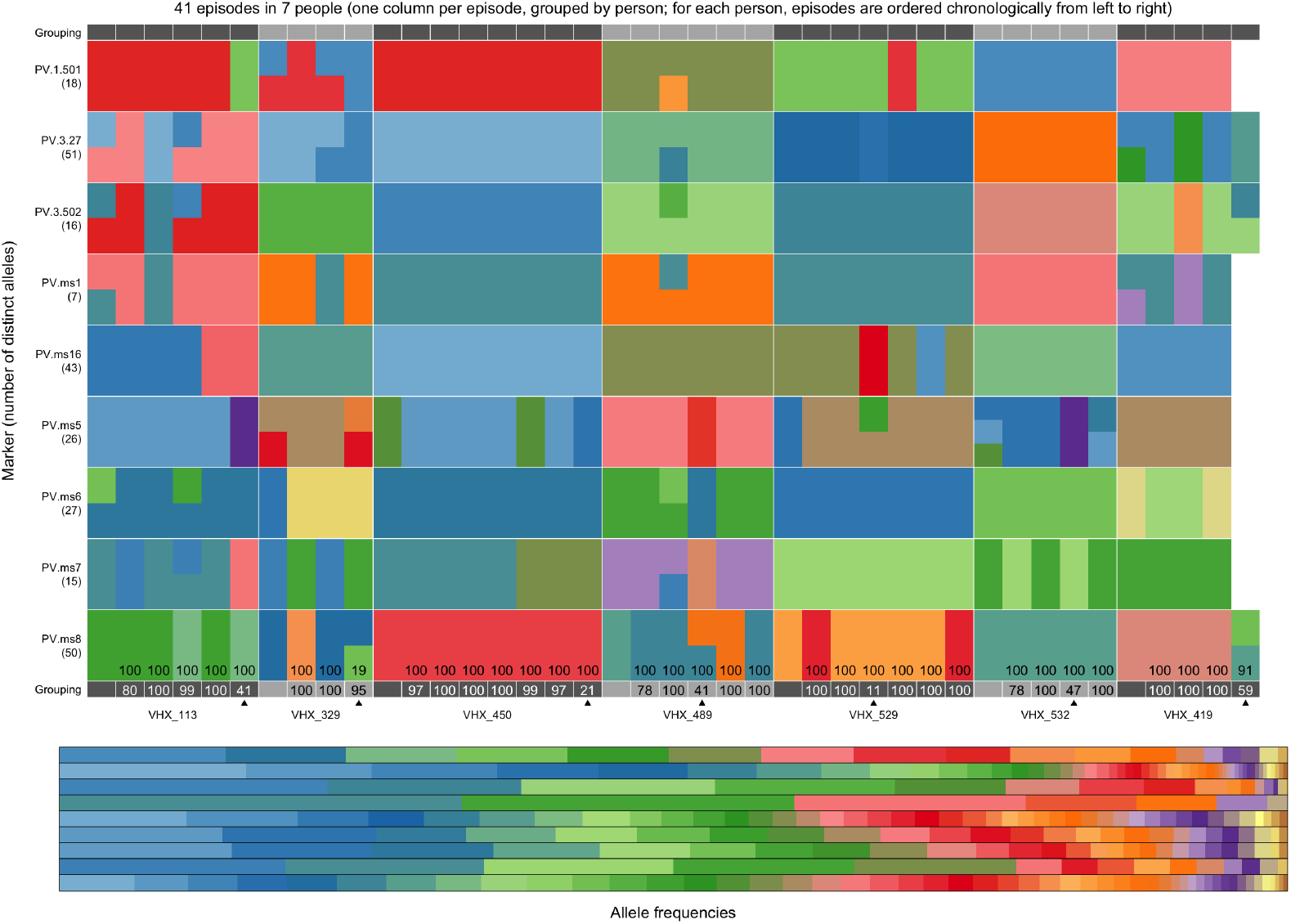
Data from study participants with recurrences whose probabilities differ by more than 0.25 for joint versus approximate joint inference with Pv3Rs. Recurrences whose probabilities differ by more than 0.25 are indicated by an arrow. The data on the sixth episode of VHX participant 419 is shown in the fifth column associated with VHX participant 419 because the first recurrence (VHX_419_2) was not successfully typed; see Figure S2. The posterior probability (×100) of relapse or recrudescence computed when data are modelled jointly is noted below that using the approximate joint approach. For all participants besides VHX 419 prior probabilities assume recurrent states are equally likely.

**Figure S9.**
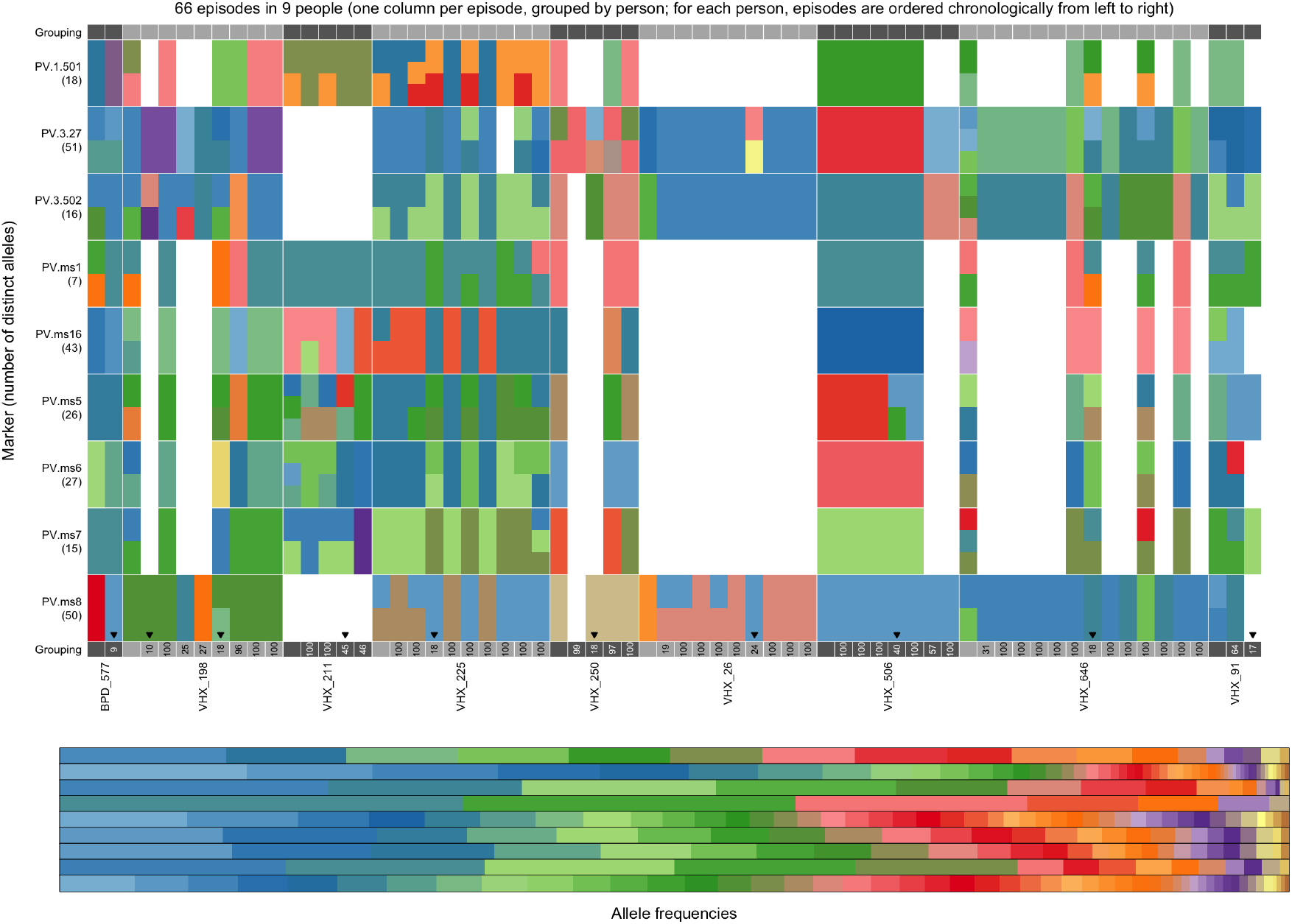
Data from study participants whose recurrences have outlying per-recurrence posterior probabilities identified using genetic relatedness and not previously, i.e., ten unlabelled red points in the top plot of Figure 2. Outliers highlighted in red in the top plot of Figure 2 are indicated by an arrow. At the bottom of each recurrences, the posterior probability (×100) of relapse plus recrudescence computed using the Pv3Rs model with a uniform prior is stated.

**Figure S10.**
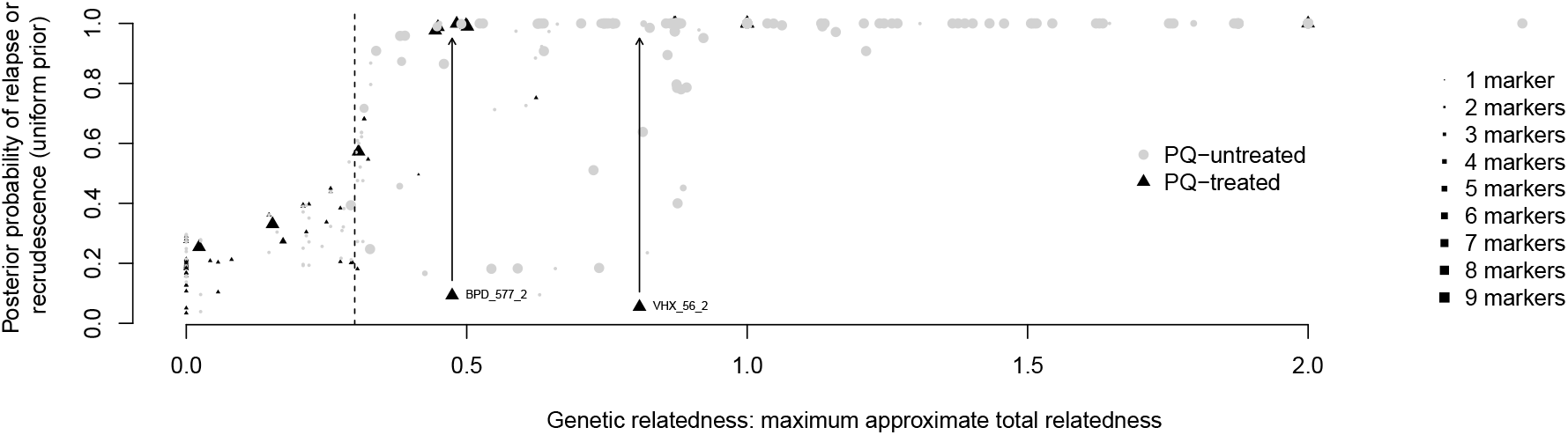
Outlier correction: an interim posterior probability of 0.990 relapse plus recrudescence is attributed to BPD_577_2 and VHX_56_2 by mapping relatedness onto probability following the sigmoid trend by eye.

**Figure S11.**
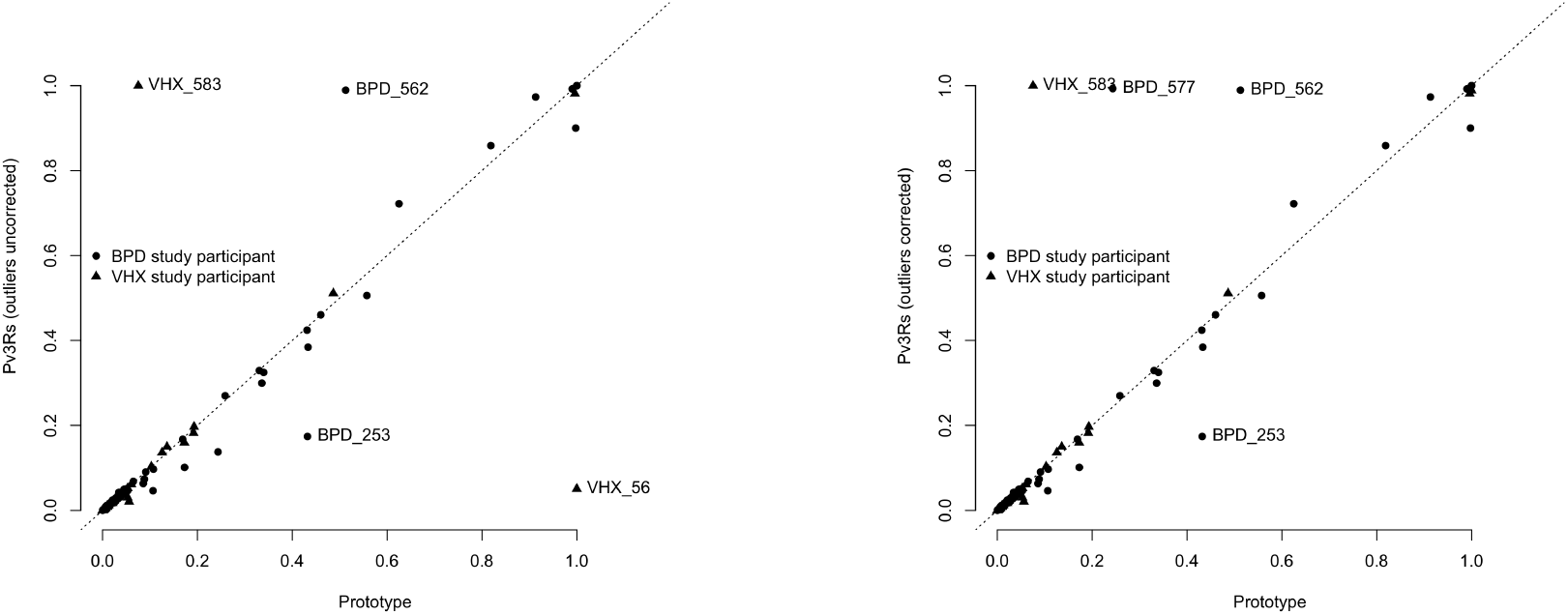
Failure rates computed using Pv3Rs-computed and prototype-computed recurrence state probabilities for PQ-treated treated study participants before (left) and after (right) the correction of outlying probabilities.

